# Utilizing Natural Language Processing and Large Language Models in the Diagnosis and Prediction of Infectious Diseases: A Systematic Review

**DOI:** 10.1101/2024.01.14.24301289

**Authors:** Mahmud Omar, Dana Brin, Benjamin Glicksberg, Eyal Klang

## Abstract

**Background:** Natural Language Processing (NLP) and Large Language Models (LLMs) hold largely untapped potential in infectious disease management. This review explores their current use and uncovers areas needing more attention.

**Methods:** This analysis followed systematic review procedures, registered with PROSPERO. We conducted a search across major databases including PubMed, Embase, Web of Science, and Scopus, up to December 2023, using keywords related to NLP, LLM, and infectious diseases. We also employed the QUADAS-2 tool for evaluating the quality and robustness of the included studies.

**Results:** Our review identified 15 studies with diverse applications of NLP in infectious disease management. Notable examples include GPT-4’s application in detecting urinary tract infections and BERTweet’s use in Lyme Disease surveillance through social media analysis. These models demonstrated effective disease monitoring and public health tracking capabilities. However, the effectiveness varied across studies. For instance, while some NLP tools showed high accuracy in pneumonia detection and high sensitivity in identifying invasive mold diseases from medical reports, others fell short in areas like bloodstream infection management.

**Conclusion:** This review highlights the yet-to-be-fully-realized promise of NLP and LLMs in infectious disease management. It calls for more exploration to fully harness AI’s capabilities, particularly in the areas of diagnosis, surveillance, predicting disease courses, and tracking epidemiological trends.

## Introduction

Artificial Intelligence (AI) marks a significant shift in streamlining medical tasks and enhancing the management of large health data, with promising implications for public health (1–3). In managing infectious diseases, a field marked by complex epidemiological and clinical challenges, AI’s potential role is notably significant (4–6).

Natural Language Processing (NLP) and generative-AI Large Language Models (LLM) are emerging as crucial yet underexplored tools in this domain (7–9). These algorithms excel at analyzing and generating text in a human-like manner, an essential capability for interpreting intricate clinical data (8–12).

The current landscape in the literature shows a mixed picture. While there are studies demonstrating AI’s potential to innovate in diagnosis and treatment planning, there is also a noticeable fragmentation in research efforts (2,4,5,10,13,14). This scenario points to both the achievements and the overlooked possibilities in AI applications, especially in the field of infectious diseases.

Our research focuses on exploring the current use of NLP and LLM in the field of infectious diseases, aiming to uncover areas where further exploration could significantly enhance diagnosis and prediction capabilities.

## Methods

### Search Strategy

This systematic review was registered with the International Prospective Register of Systematic Reviews - PROSPERO (Registration code: CRD42023494253). We adhered to the Preferred Reporting Items for Systematic Reviews and Meta-Analyses (PRISMA) guidelines (15,16).

A systematic search was conducted across key databases: PubMed, Embase, Web of Science, and Scopus, up until December 2023. We complemented the search via reference screening for any additional papers.

We aimed to identify original research articles that investigated the application of NLP and LLM in the diagnosis or prediction of infectious diseases.

The search utilized a combination of keywords including “infectious diseases”, “infection”, “LLM”, “Large Language Mode”, “AI”, “Artificial Intelligence”, “natural language processing”, and “NLP”. Specific search strings for each database are detailed in the supplementary materials.

### Study Selection

We included original research articles that focused on the application of NLP and LLM in diagnosing or predicting infectious diseases.

Studies were selected if they provided data for assessing the performance metrics of AI models, such as area under the curve, accuracy, sensitivity, and specificity.

We excluded review papers, case reports, conference abstracts, editorials, preprints, and studies not conducted in English. We also excluded studies employing classic machine-learning techniques unrelated to NLP.

### Data Extraction

Two independent reviewers extracted relevant information using a standardized form. Data points included the first author’s name, year of publication, study design, sample size, specific conversational NLP techniques used, dataset details for model training and validation, performance metrics, and key findings. Discrepancies between reviewers were resolved through discussion, and a third reviewer was consulted when necessary.

### Risk of Bias

To evaluate the quality and robustness of the methodologies in the included studies, the QUADAS-2 (Quality Assessment of Diagnostic Accuracy Studies-2) tool was used (17).

## Results

### Natural Language Processing (NLP) and Generative AI Models

Natural Language Processing (NLP), a vital part of AI, enables machines to understand and interpret human language text (7). Within this domain, generative AI models, especially Large Language Models (LLMs), are gaining prominence (10,18). These models are designed to generate text that closely resembles human writing, providing valuable insights and applications.

Among the notable LLMs are Bidirectional Encoder Representations from Transformers (BERT) and Generative Pre-trained Transformers (GPT). BERT is particularly effective at grasping the context within language, allowing for more accurate interpretations of text (19). On the other hand, GPT stands out for its ability to produce text that is remarkably similar to human-generated content, a feature that has broad applications in various fields including healthcare (20).

Together, NLP and these advanced generative models are reshaping how we interact with and utilize language data in the AI sphere, opening new avenues for research and application (**Figures 1 and 2**).

**Figure 1:**
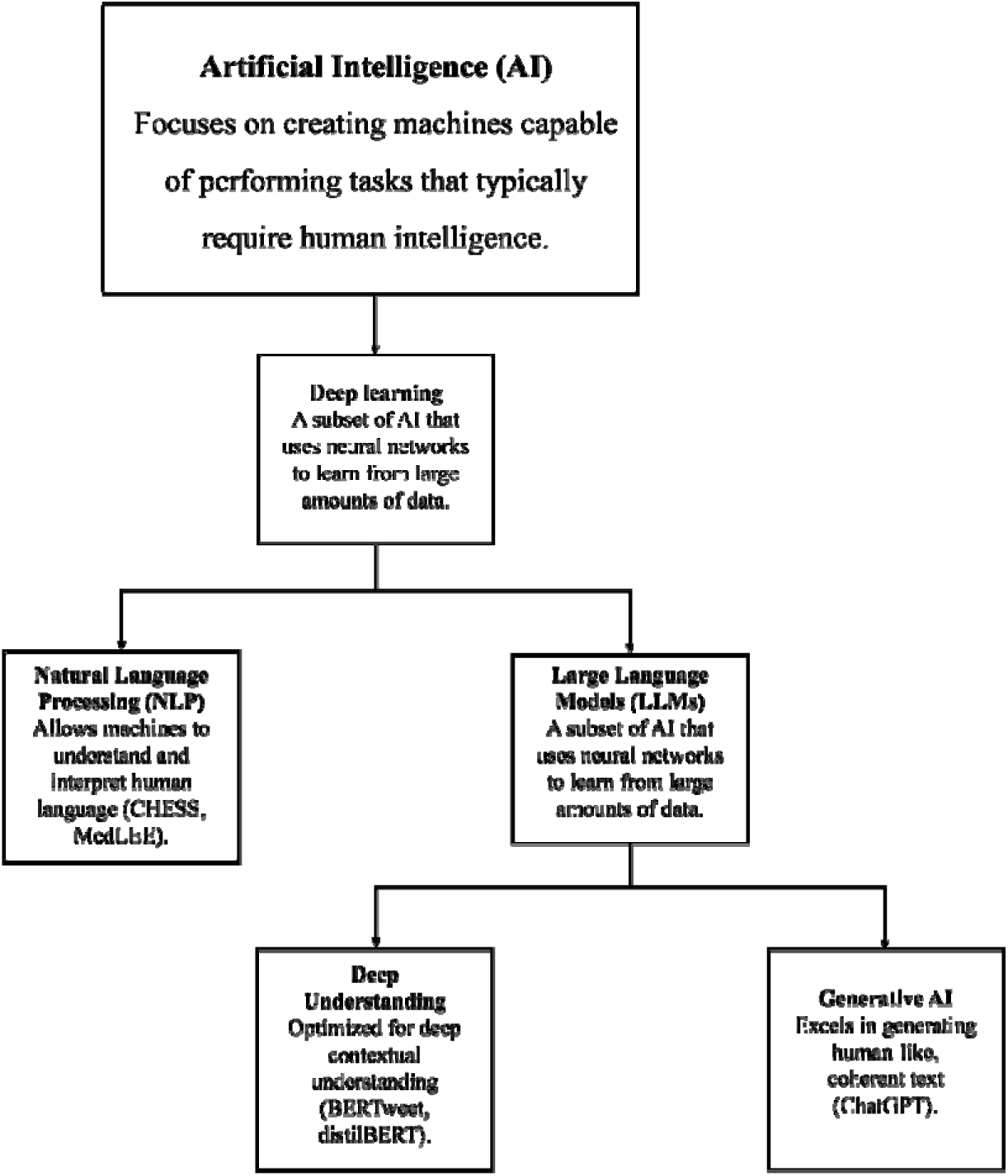
A Framework of AI in Language Processing.

**Figure 2:**
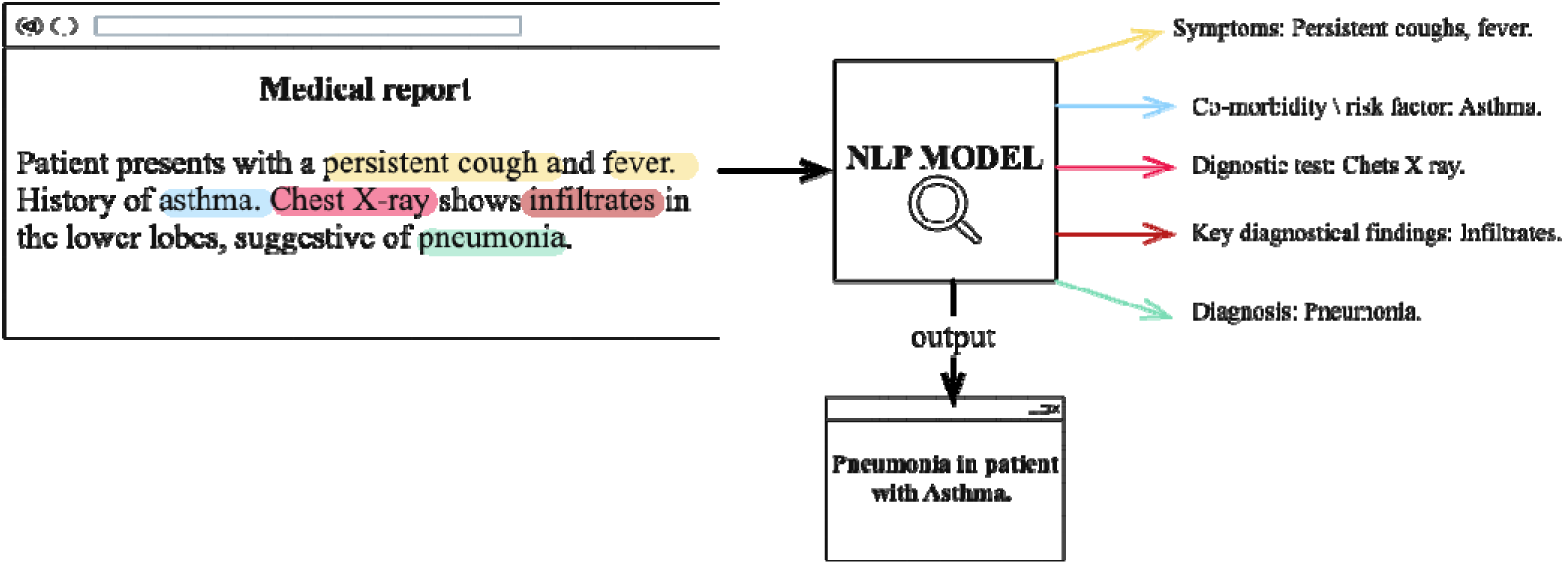
Decoding Clinical Narratives: An NLP Model’s Path from Text to Diagnosis.

### Search Results and Study Selection

Our search yielded a total of 432 articles. Specifically, the search resulted in 177 articles from PubMed, 52 from Embase, 68 from Web of Science, and 135 from Scopus.

After the removal of 143 duplicates, the screening process found 13 studies that met our inclusion and exclusion criteria (19,21–32). We identified two additional studies via reference screening (33,34).

These studies varied in their disease focus, underscoring the diverse applications of NLP and LLM in the field of infectious diseases.

The process of study selection and the screening methodology are detailed in the PRISMA flow chart (**Figure 3**).

**Figure 3:**
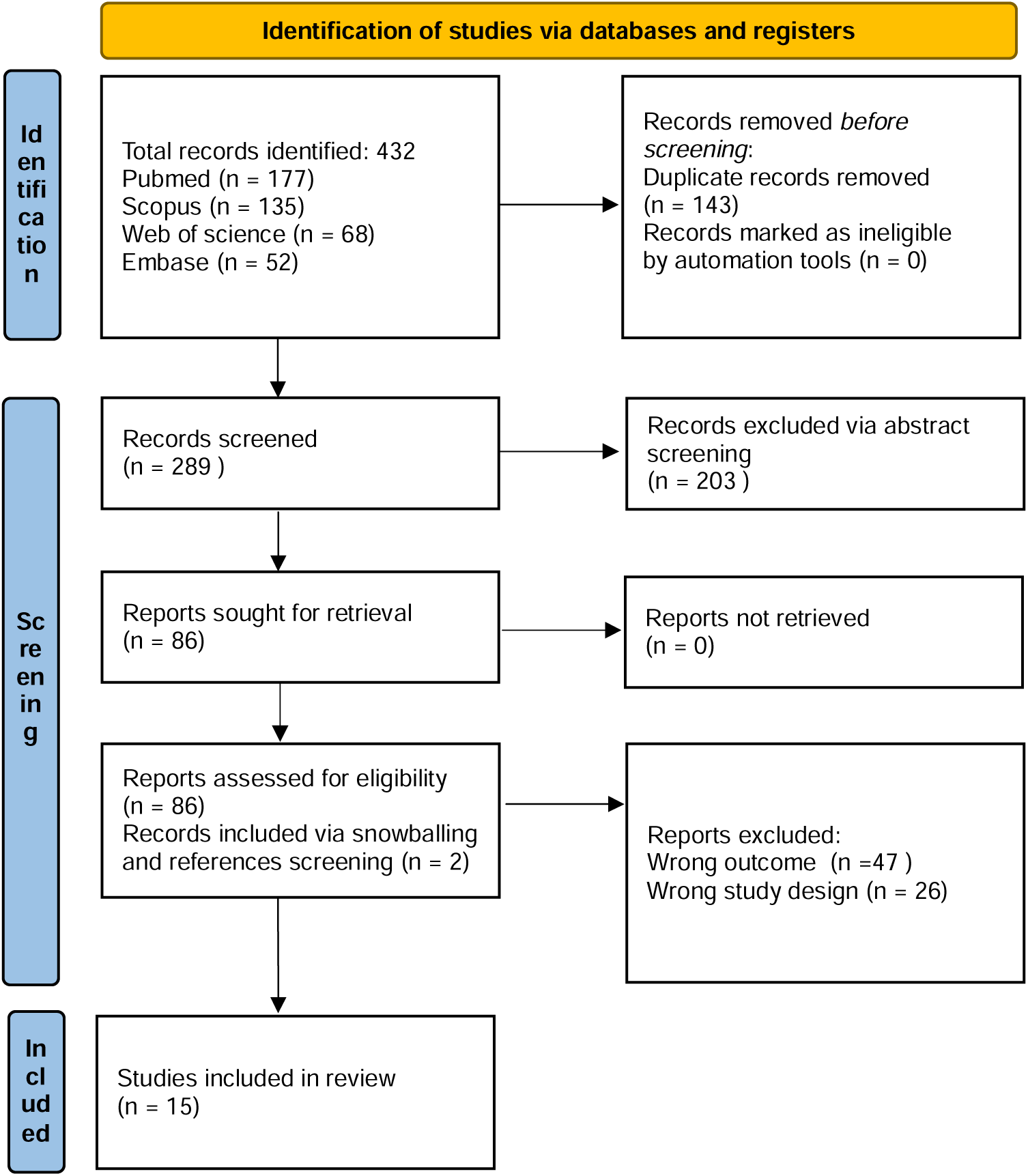
PRISMA flowchart.

### Risk of Bias

Six studies demonstrated a low risk of bias across all domains (21,31–35). Overall, ten studies were evaluated as having low risk (21–23,26,28,31–35), four showed high risk (19,24,27,29), and one lacked sufficient data for a conclusive risk assessment (30).

Generally, the studies indicated a predominantly low risk of bias, suggesting reliable and valid results (**Figures 4-5**). Concerning applicability, most of the studies demonstrated a low risk. However, specific concerns were raised in a few studies, particularly regarding their study populations. Several studies used specialized populations, such as patients with hematologic neoplasms or data derived from specific databases, which may limit generalizability (25–29,35).

**Figure 4:**
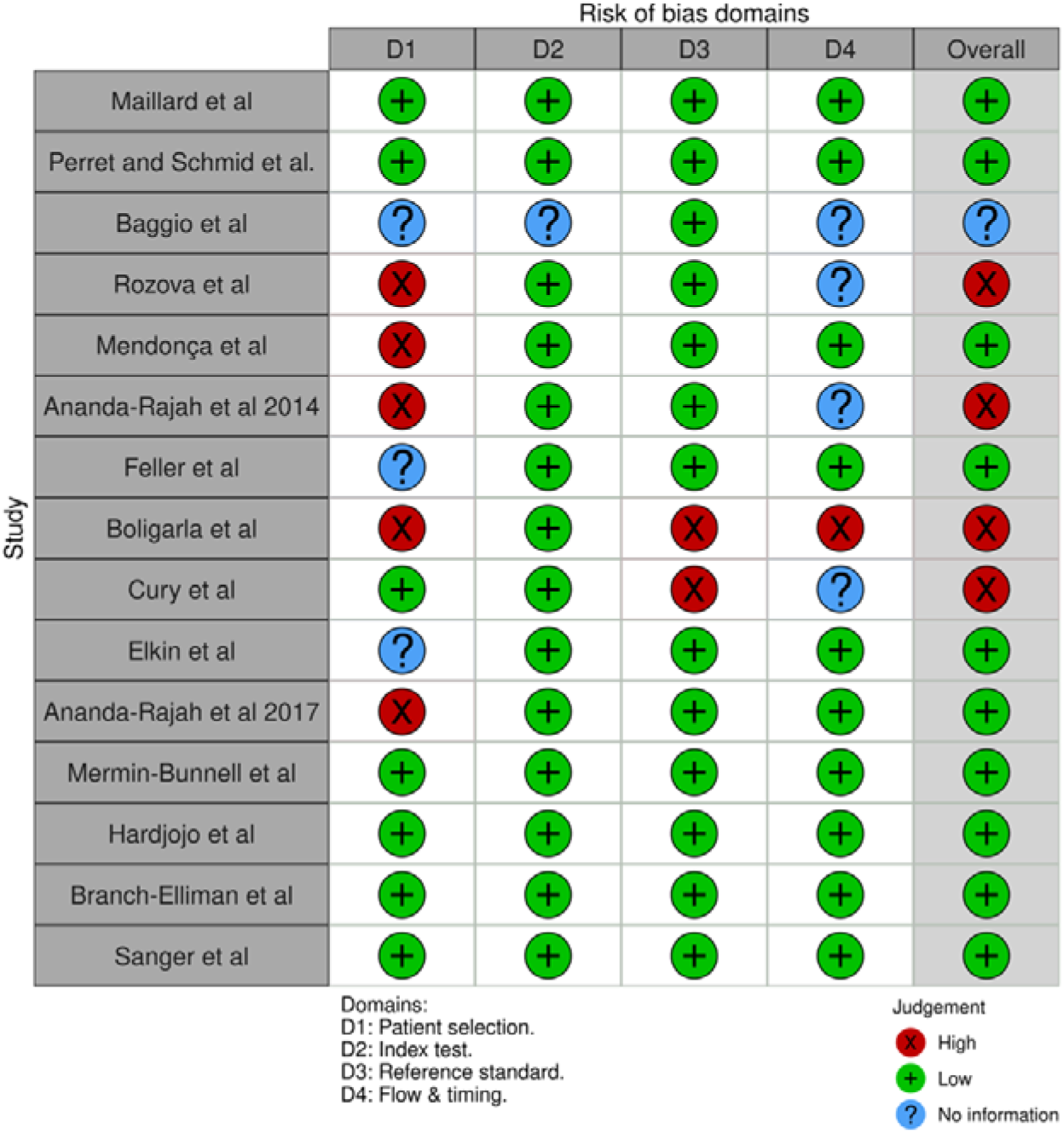
Distribution of Risk of Bias Concerns Across Individual Domains.

**Figure 5:**
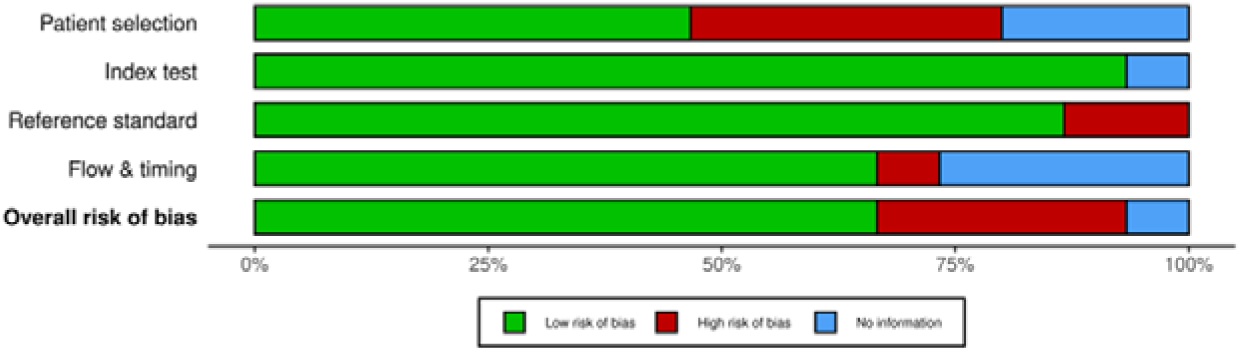
Cumulative Assessment of Overall Risk of Bias Concerns Across All Included Studies.

### Overview of Included Studies

The included studies were published between 2005 and 2023 and present a comprehensive view of the evolving use of NLP and LLM in diagnosing and managing infectious diseases. The studies focused on various diseases, including fungal infections in immunocompromised patients, respiratory infections in newborns, HIV, Lyme disease, and COVID-19. These studies utilized different data sources such as radiology and histopathology reports, clinical data from medical files, and social media posts (e.g., tweets). Notably, two studies tested ChatGPT-4, two studies utilized BERT, while others employed a variety of NLP models. This showcases the evolving landscape of AI applications in infectious diseases.

Notably, Maillard et al. and Perret and Schmid et al. utilized GPT-4 to analyze bloodstream infections and catheter-associated urinary tract infections, respectively (31,35). They demonstrated the potential of AI in clinical decision-making despite certain limitations.

Baggio et al. and Ananda-Rajah et al. employed machine learning-based NLP to enhance the surveillance of invasive mold diseases (22,27,30).

Additionally, studies like Feller et al. and Boligarla et al. explored innovative applications of NLP in risk assessment and disease surveillance using diverse data sources, including clinical notes and social media (25,26).

These studies collectively underscore the significant advancements and potential of NLP in infectious disease research, while also highlighting areas for improvement and further exploration (**Table 1**).

**Table 1:**
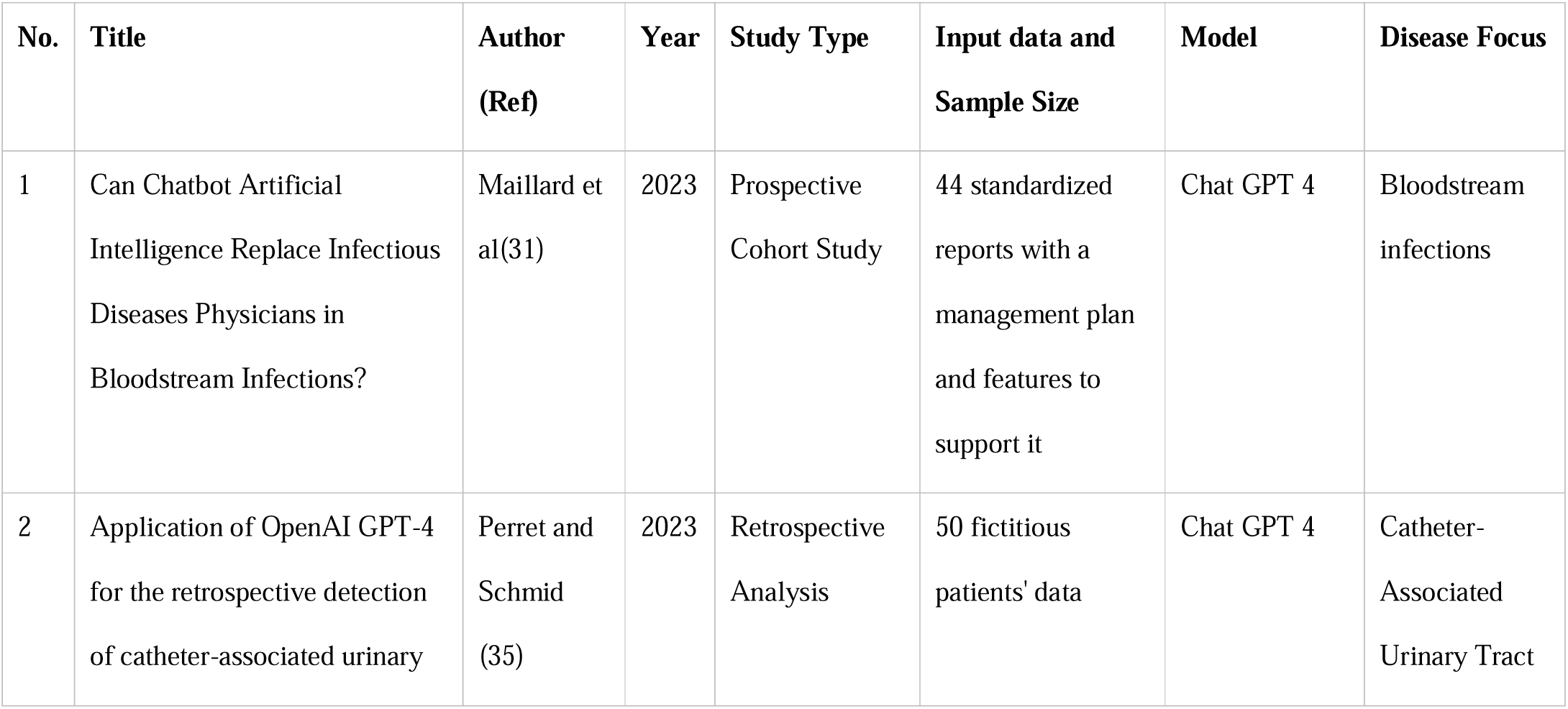
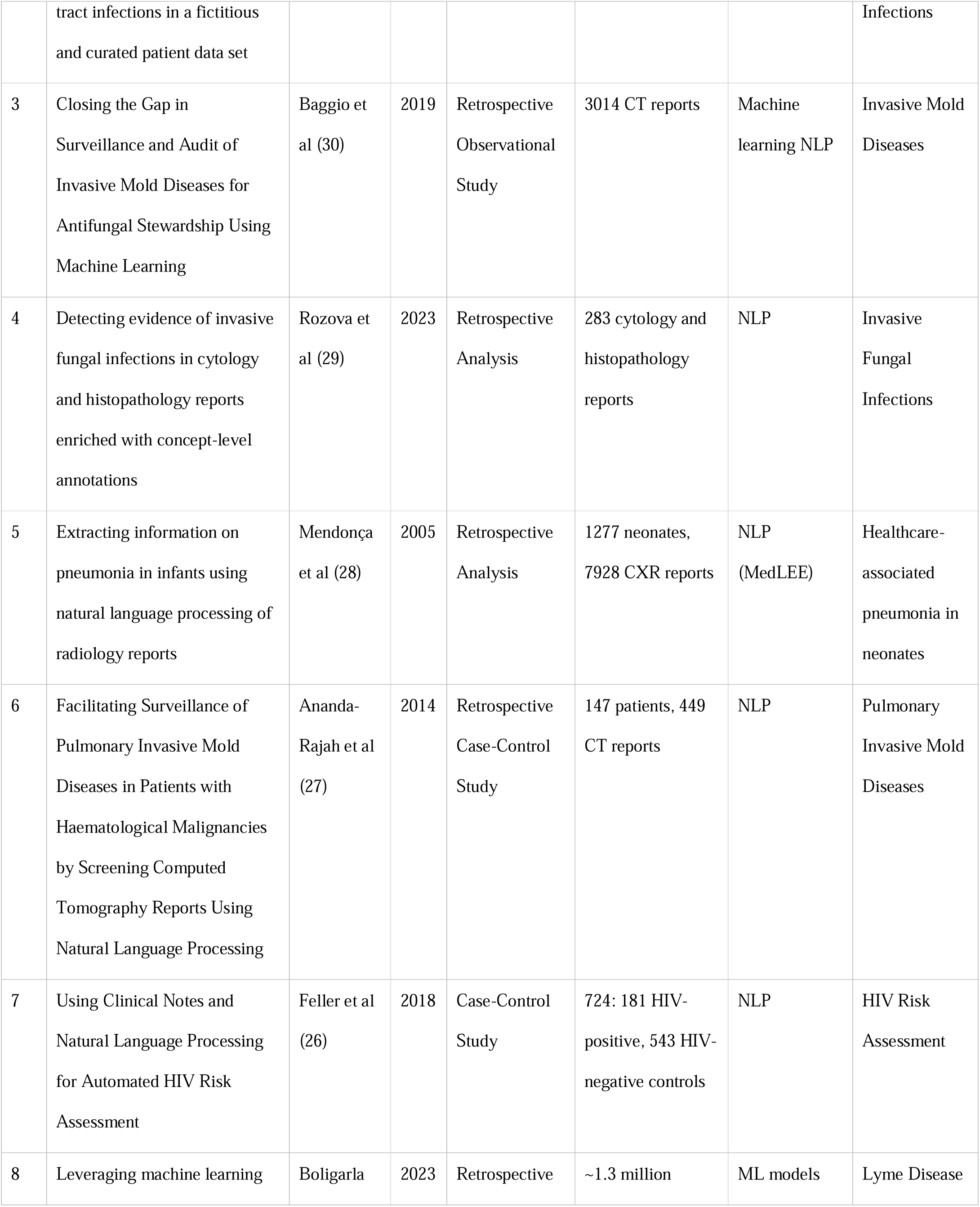
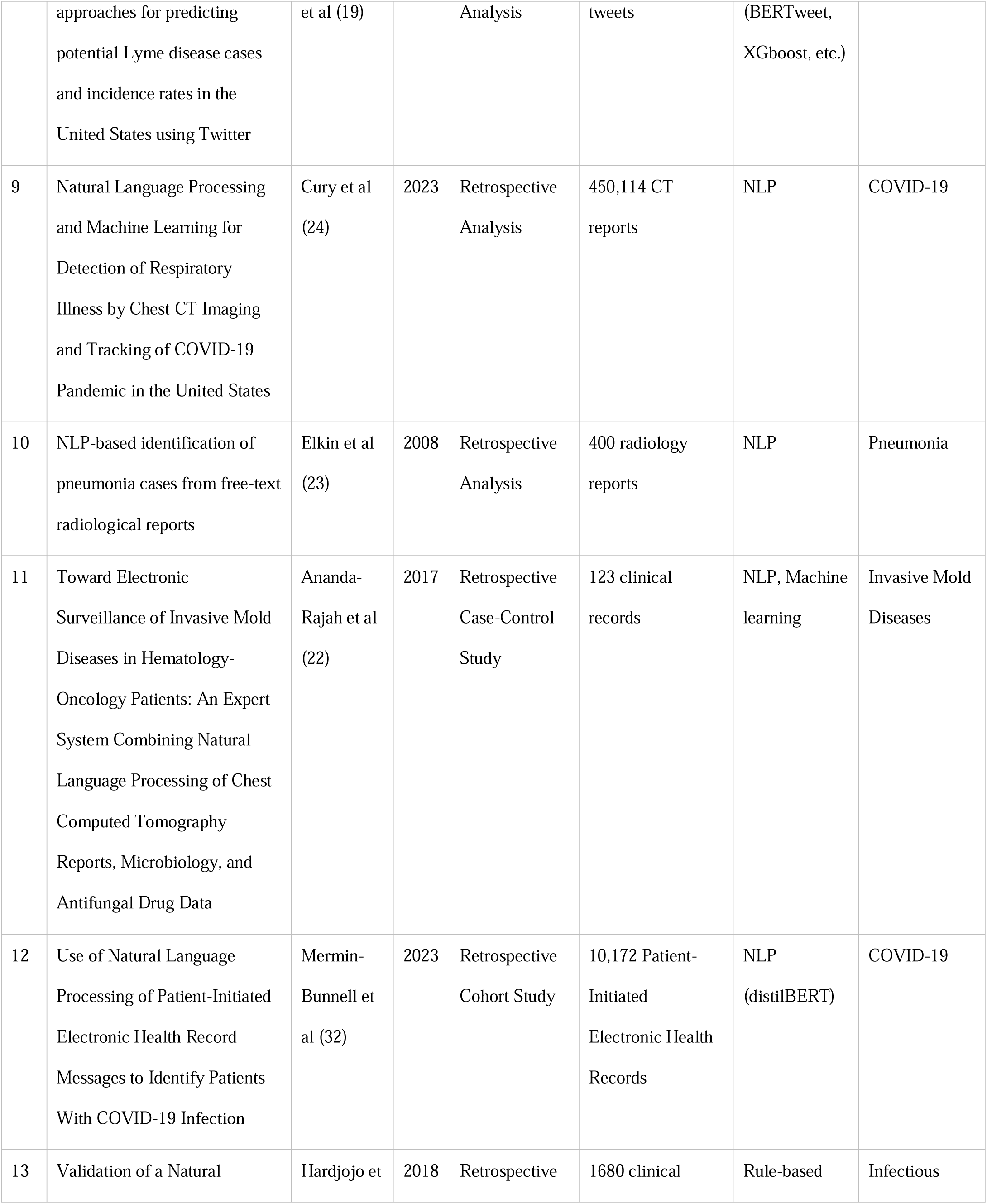
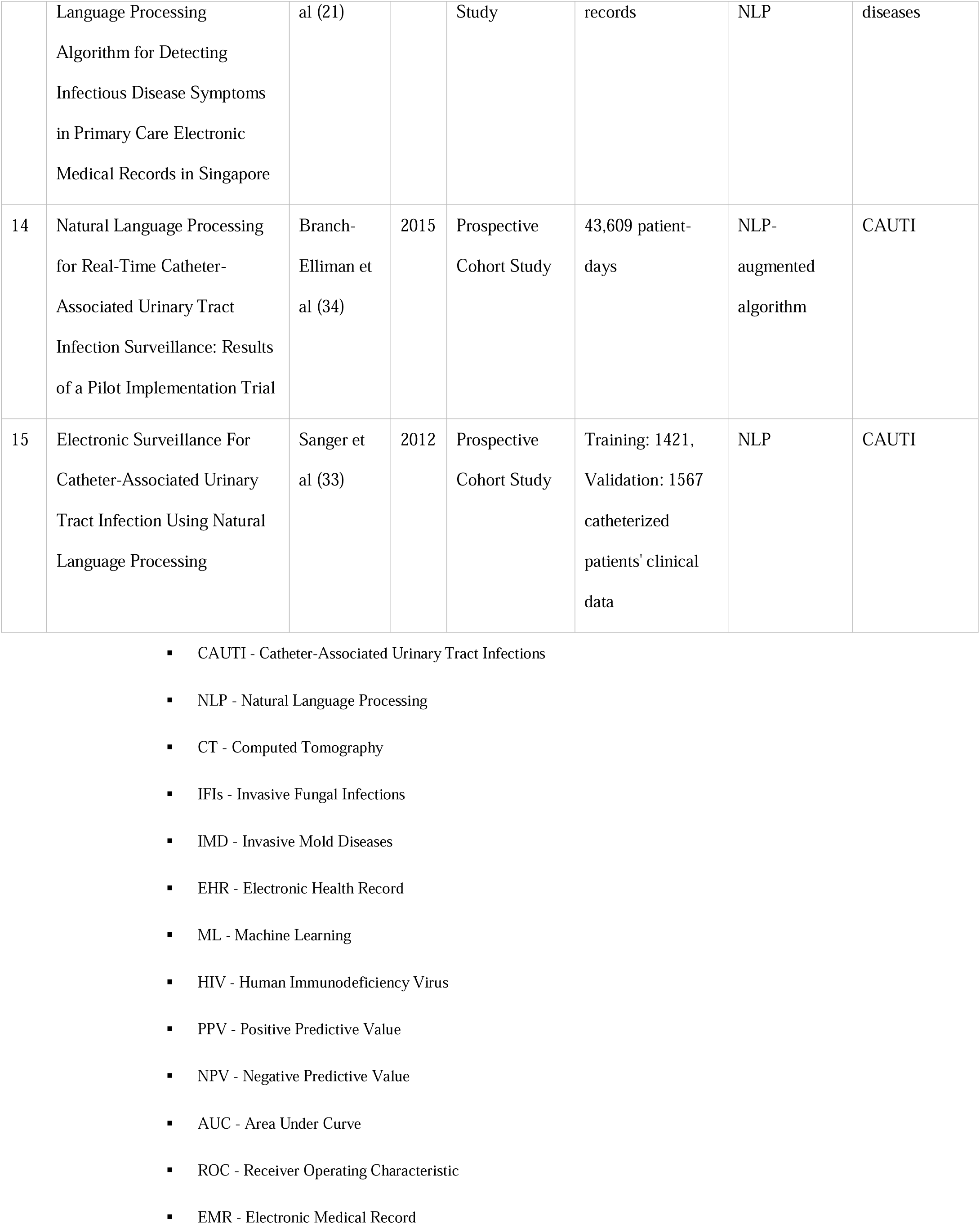
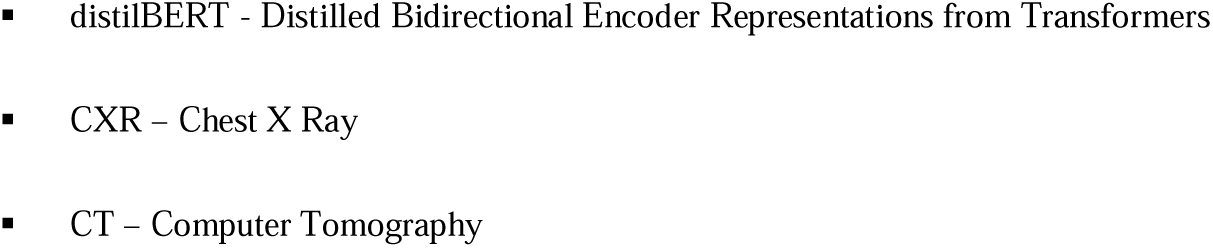
Summary of the included studies.

### Narrative Synthesis of AI and NLP in Infectious Diseases

#### Diagnosis and patient management

In comparing the effectiveness of NLP and LLM in infectious disease management, the results show varied outcomes. GPT-4, as used in Maillard et al.’s and Perret and Schmid et al.’s studies (31,35), demonstrated high accuracy in catheter-associated urinary tract infections (CAUTI) detection. However, the model showed moderated effectiveness in bloodstream infection management.

NLP algorithms in the studies by Branch-Elliman et al. and Sanger et al. showed high sensitivity and specificity for CAUTI detection (33,34). This indicates their strong potential for precise medical surveillance and diagnosis.

#### NLP in Disease Monitoring from Medical Reports

NLP demonstrated high effectiveness in pneumonia and invasive mold disease detection. For instance, the MedLEE model achieved up to 99% specificity in pneumonia detection (28). In Baggio et al, an NLP classifier showed 91% sensitivity in identifying invasive mold diseases from CT reports (30). These results underline NLP’s promising role in disease surveillance.

#### Public Health Surveillance

Several studies revealed NLP’s effectiveness in health monitoring.

For example, BERTweet model was used for Lyme Disease detection through social media tweets (25). BERTweet, analyzing 1.3 million tweets, reached 90% accuracy.

The eCOV model was used for COVID-19 electronic health records (EHR) messages (32). eCOV’s classification scored a 94% macro F1.

#### Prediction and Risk Assessment

NLP models have shown advancements in infectious disease risk assessment.

For example, an HIV risk assessment model achieved an F1 score of 0.74, demonstrating NLP’s capability in extracting vital clinical data (29).

Another model, CHESS, used in detecting infectious disease symptoms in primary care records, achieved high precision and recall (21).

These results highlights NLP’s efficiency in predictive analytics from varied clinical data sources.

### Key Summary of LLM and NLP in Infectious Disease Research

1. **Public Health Surveillance**: Tools like BERTweet are effective in tracking disease epidemiology through social media analysis.
2. **Diagnostic Accuracy**: NLP shows high accuracy in disease detection from medical reports. This could be helpful for incidence monitoring and for aiding diagnoses (**Table 2**).
3. **Clinical Applications**: NLP and LLM have versatile use cases: disease monitoring, risk assessment, pandemic prevention, and prediction, and aiding clinical decisions (**Table 3**).

## Discussion

The integration of NLP and LLM in the management of infectious diseases holds the promise for a paradigm shift in medical diagnostics and public health surveillance (20,36–39). Our systematic review reveals a landscape where the potential of these technologies is only beginning to be tapped.

**Table 2:** Performance Metrics and Key Findings.

| Author (Ref) | Model | Clinical Task | Performance Metrics | Main Findings |
| --- | --- | --- | --- | --- |
| Maillard et al(31) | ChatGPT-4 | Bloodstream<br>Infection<br>Diagnosis and<br>Management | Diagnostic accuracy:<br>59%, Empiric therapy:<br>64% | ChatGPT-4 had detailed responses<br>but suboptimal and even hazardous<br>management plans in complex<br>cases. |
| Perret and<br>Schmid (35) | ChatGPT-4 | CAUTI Detection | Sensitivity: 91%,<br>Specificity: 92% | High accuracy in CAUTI detection<br>from patient data. |
| Baggio et al (30) | NLP (ML-based) | IMD<br>Identification<br>from CT Reports | Sensitivity: 91%,<br>Specificity: 79%, AUC:<br>0.92 | Effective identification of IMD<br>cases, aiding antifungal<br>stewardship. |
| Rozova et al (29) | NLP | IFI Diagnosis<br>from Cytology<br>and<br>Histopathology | Micro-F1: 0.84 (dev),<br>0.83 (test) | Efficient IFI identification using<br>concept annotations. |

|  |  | Reports |  |  |
| --- | --- | --- | --- | --- |
| Mendonça et al (28) | MedLEE | Neonatal Pneumonia Detection | Sensitivity: 71%, Specificity: 99%, PPV: 7.5% | Feasible for identifying pneumonia, but low PPV due to broad definition. |
| Ananda-Rajah et al (27) | NLP (ML-based) | IMD Detection in Hematological Malignancies | Sensitivity: 91%, Specificity: 79%, AUC: 0.92 | Effective in IMD surveillance for hematological malignancies. |
| Feller et al (26) | NLP | HIV Risk Assessment | F1 score: 0.74 | Improved HIV risk prediction incorporating clinical keywords. |
| Boligarla et al (19) | BERTweet and other ML models | Lyme Disease Case Prediction | Accuracy and F1-score: 90% | Effective in identifying Lyme-related tweets, strong correlation with disease counts. |
| Cury et al (24) | NLP | COVID-19 Respiratory Illness Detection | Correlation coefficients: Viral Pneumonia ($r^2=0.13$), Imaging Findings ($r^2=0.66$), COVID NLP ($r^2=0.82$) | High correlation with official COVID-19 case counts, early detection capability. |
| Elkin et al (23) | NLP | Pneumonia Detection from Radiological Reports | Sensitivity: 100%, Specificity: 90.3-97.99%, PPV: 70-97.44% | High accuracy for pneumonia identification. |
| Ananda-Rajah et al (22) | NLP, ML | IMD Surveillance in Hematology-Oncology Patients | Specificity: 74.6%, Sensitivity: 98.4%, ROC: 92.8% | Combined NLP and data sources for improved IMD prediction. |
| Mermin-Bunnell et al (32) | distilBERT | COVID-19 Case Classification from EHR Messages | Macro F1: 94%, Sensitivity: 85-100% | High accuracy in classifying COVID-19 related EHR messages. |
| Hardjojo et al (21) | CHESS | Infectious Disease Symptom Detection in EMRs | Precision: 96.0%, Recall: 93.1% | High precision and recall in identifying infectious disease symptoms in EMRs. |
| Branch-Elliman et al (34) | NLP-augmented | Indwelling Urinary Catheter Days and CAUTI Identification | PPV: 54.2%, Sensitivity: 65% | Identified more catheter days, variation in performance between unit types. |
| Sanger et al (33) | NLP | CAUTI Detection from Clinical Data | Sensitivity: 97.1%, Specificity: 94.5%, PPV: 66.7%, NPV: 99.6% | High sensitivity and specificity, ideal as a screening tool. |

- AI - Artificial Intelligence - NLP - Natural Language Processing - CAUTI - Catheter-Associated Urinary Tract Infections - IMD - Invasive Mold Diseases - IFIs - Invasive Fungal Infections - AUC - Area Under Curve - PPV - Positive Predictive Value - NPV - Negative Predictive Value - EHR - Electronic Health Record - HIV - Human Immunodeficiency Virus - ROC - Receiver Operating Characteristic - CT - Computed Tomography - IUCD - Indwelling Urinary Catheter Days

**Table 3:**
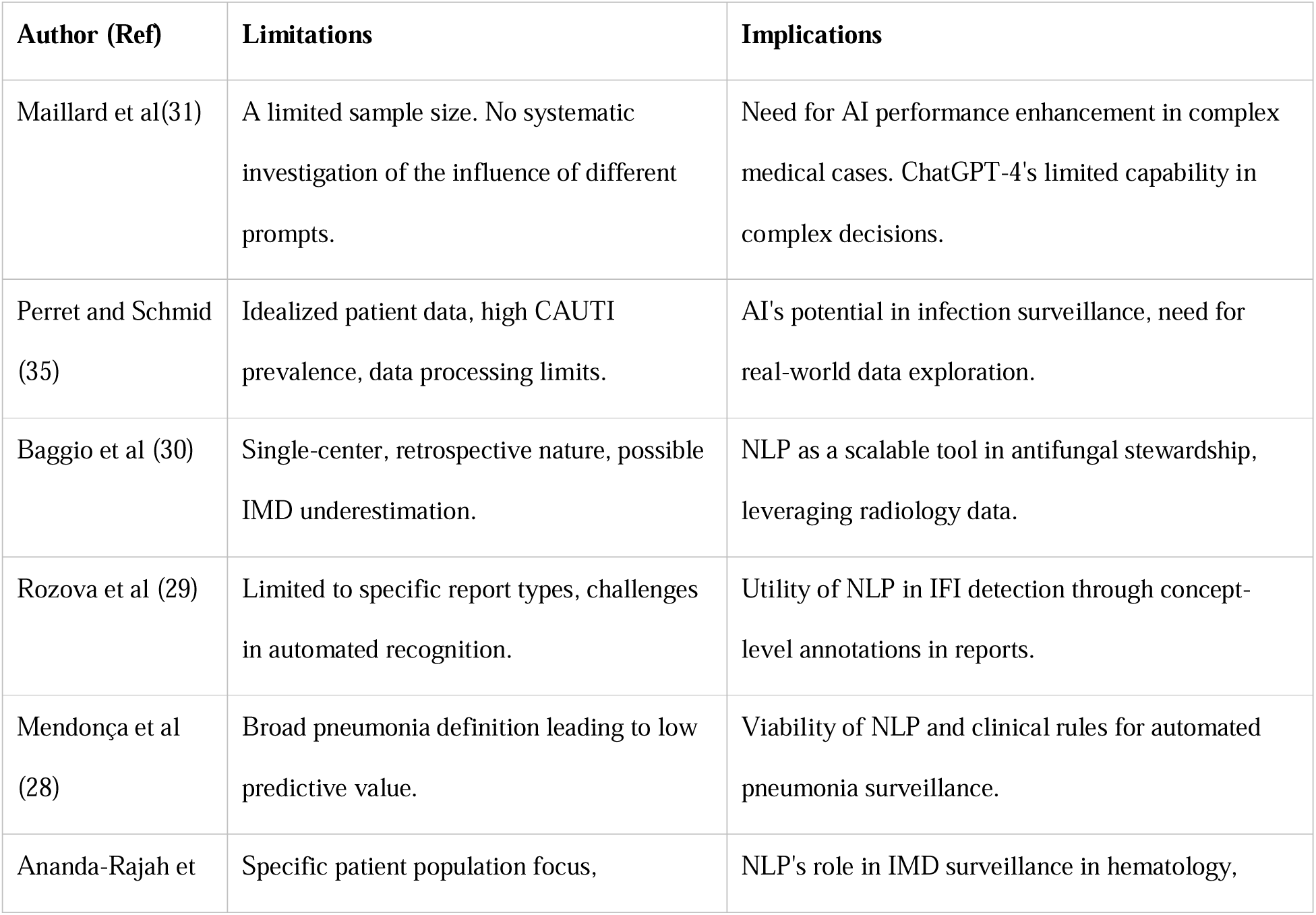
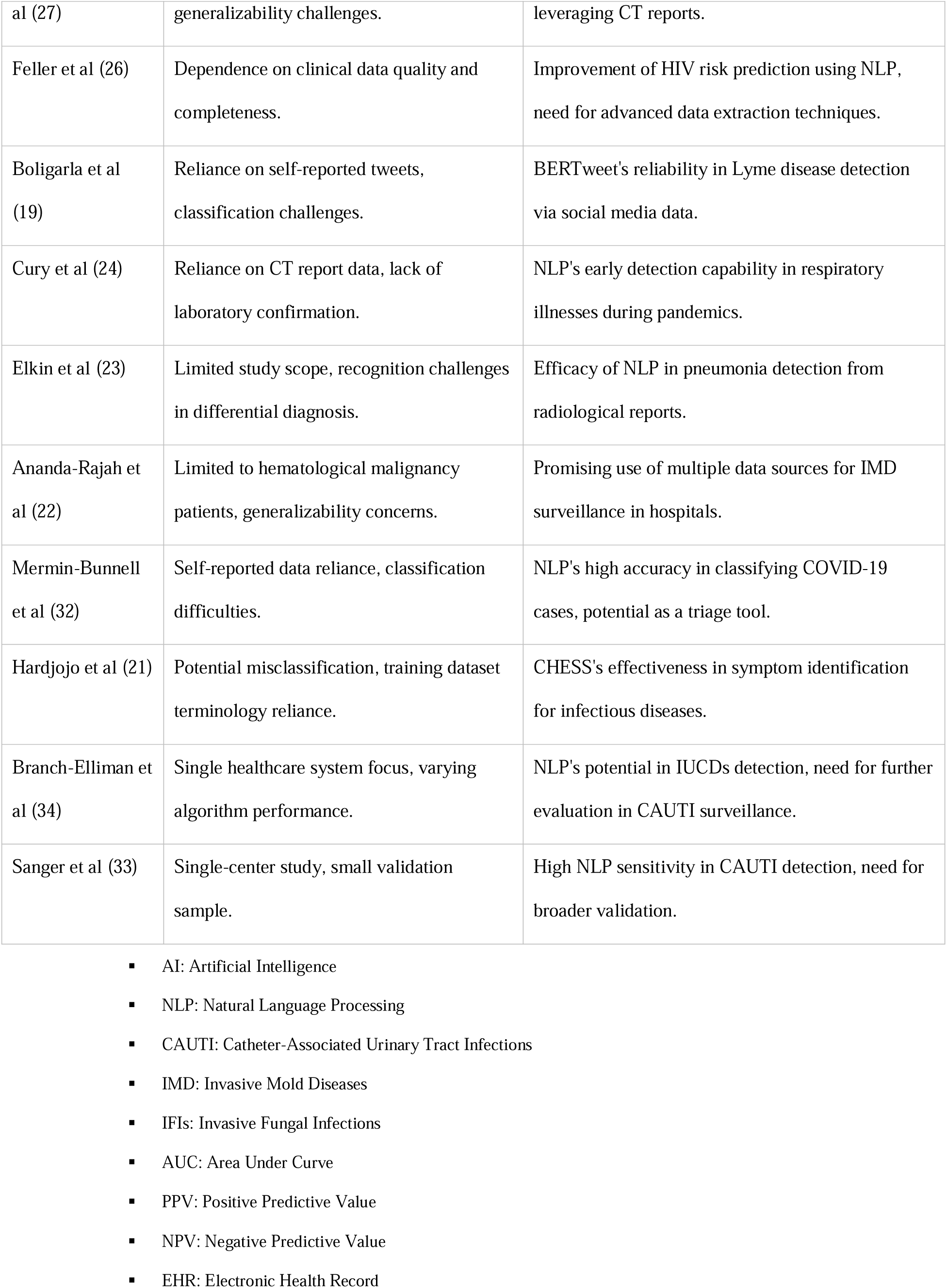
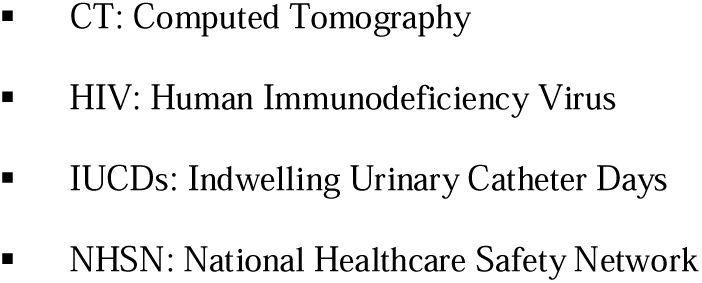
Limitations and Implications.

Firstly, the implementation of NLP and LLM in infectious diseases showcases a leap forward in medical technology’s ability to process and interpret complex clinical data (1,2,36,37). The precision in disease detection, as illustrated by studies on CAUTI and pneumonia, underscores the capability of AI to augment human expertise in diagnostic processes (23,24,33–35). However, the variation in effectiveness across different infectious diseases highlights the nuanced nature of AI applications in healthcare. For instance, a study by Wilhelm et al. on various Conversational AI models across different medical specialties like ophthalmology, orthopedics, and dermatology, found significant differences in the quality and safety of the medical advice provided (40). Furthermore, research by Maillard and colleagues revealed that GPT-4 occasionally produces incorrect and, at times, detrimental medical guidance and treatment strategies (31). It is not a one-size-fits-all solution but requires tailored approaches depending on the disease context. Secondly, the use of AI in public health surveillance, as evidenced by the BERTweet model for Lyme Disease and the eCOV model for COVID-19, is a breakthrough (19,24,32). These tools demonstrate how AI can transform unstructured data from diverse sources into actionable insights, a critical advancement for early outbreak detection and response.

Thirdly, the evolution of AI in risk assessment and predictive analytics marks a significant stride. Models like the HIV risk assessment tool and CHESS show that AI can efficiently sift through vast clinical data, extracting crucial information that can inform patient care and disease management (21,26).

However, this promising landscape is not without its challenges. The variability in study methodologies and the limited focus of some studies on specific patient populations or diseases indicate that generalizability remains a concern. The dependency on the quality and completeness of clinical data for AI models underscores the need for robust data infrastructure in healthcare settings. For example, a lack of contextual awareness and inherent biases may sometimes hinder AI safe deployment in clinical settings, as reported by Schwartz et al (5).

Furthermore, the nascent stage of LLM application in infectious diseases, as seen in the limited number of studies, suggests that we are only scratching the surface of what these technologies can achieve (10,20,36).

### Limitations

Our study faces several limitations. The emerging nature of LLM in healthcare research means that there are relatively few studies specifically focusing on this technology, limiting our ability to draw comprehensive conclusions. The immense potential of LLM necessitates more extensive research to fully understand its capabilities and limitations. Additionally, the predominance of retrospective studies in our review indicates a need for more prospective studies to validate the findings (41). Lastly, the diverse methodologies and focus areas of the included studies prevented us from performing a meta-analysis, highlighting the need for more standardized research approaches in this field.

### Conclusion

This review highlights the yet-to-be-fully-realized promise of NLP and LLMs in infectious disease management. It calls for more exploration to fully harness AI’s capabilities, particularly in the areas of diagnosis, surveillance, predicting disease courses, and tracking epidemiological trends.

## Data Availability

All data produced in the present study are available upon reasonable request to the authors

## Acknowledgment

none

## Supplementary materials

**Appendix** – Screening Boolean strings for the different databases.

**Pubmed**

(”large language models” OR “LLM” OR “ChatGPT” OR “GPT-3” OR “GPT-4” OR “natural language processing” OR “NLP”) AND (”infectious diseases” OR “bacterial infections” OR “viral infections” OR “fungal infections” OR “parasitic infections”)

**Scopus**

(TITLE-ABS-KEY(”large language models” OR “LLM” OR “ChatGPT” OR “GPT-3” OR “GPT-4” OR “natural language processing” OR “NLP”) AND TITLE-ABS-KEY(”infectious diseases” OR “bacterial infections” OR “viral infections” OR “fungal infections” OR “parasitic infections”))

**Web of science**

(TS=(”large language models” OR “LLM” OR “ChatGPT” OR “GPT-3” OR “GPT-4” OR “natural language processing” OR “NLP”)) AND TS=(”infectious diseases” OR “bacterial infections” OR “viral infections” OR “fungal infections” OR “parasitic infections”)

**Embase**

(’large language models’:ab,ti OR ’LLM’:ab,ti OR ’ChatGPT’:ab,ti OR ’GPT-3’:ab,ti OR ’GPT-4’:ab,ti OR ’natural language processing’:ab,ti OR ’NLP’:ab,ti) AND (’infectious diseases’:ab,ti OR ’bacterial infections’:ab,ti OR ’viral infections’:ab,ti OR ’fungal infections’:ab,ti OR ’parasitic infections’:ab,ti)

